# Integrating HiTOP and RDoC Frameworks Part I: Genetic Architecture of Externalizing and Internalizing Psychopathology

**DOI:** 10.1101/2024.04.06.24305166

**Authors:** Christal N. Davis, Yousef Khan, Sylvanus Toikumo, Zeal Jinwala, Dorret I. Boomsma, Daniel F. Levey, Joel Gelernter, Rachel L. Kember, Henry R. Kranzler

## Abstract

**Background:** There is considerable comorbidity between externalizing (EXT) and internalizing (INT) psychopathology. Understanding the shared genetic underpinnings of these spectra is crucial for advancing knowledge of their biological bases and informing empirical models like the Research Domain Criteria (RDoC) and Hierarchical Taxonomy of Psychopathology (HiTOP).

**Methods:** We applied genomic structural equation modeling to summary statistics from 16 EXT and INT traits in European-ancestry individuals (n = 16,400 to 1,074,629). Traits included clinical (e.g., major depressive disorder, alcohol use disorder) and subclinical measures (e.g., risk tolerance, irritability). We tested five confirmatory factor models to identify the best fitting and most parsimonious genetic architecture and then conducted multivariate genome-wide association studies (GWAS) of the resulting latent factors.

**Results:** A two-factor correlated model, representing EXT and INT spectra, provided the best fit to the data. There was a moderate genetic correlation between EXT and INT (r = 0.37, SE = 0.02), with bivariate causal mixture models showing extensive overlap in causal variants across the two spectra (94.64%, SE = 3.27). Multivariate GWAS identified 409 lead genetic variants for EXT, 85 for INT, and 256 for the shared traits.

**Conclusions:** The shared genetic liabilities for EXT and INT identified here help to characterize the genetic architecture underlying these frequently comorbid forms of psychopathology. The findings provide a framework for future research aimed at understanding the shared and distinct biological mechanisms underlying psychopathology, which will help to refine psychiatric classification systems and potentially inform treatment approaches.

## Introduction

Traditional categorical classifications of psychopathology are limited by a rudimentary understanding of the underlying biology. Psychiatric disorders co-occur more often than expected by chance (Kessler, Chiu, Demler, & Walters, 2005; Kessler et al., 1994), suggesting the presence of shared genetic underpinnings across diagnostic categories (Lee, Feng, & Smoller, 2021). Additionally, current diagnostic classifications rely on arbitrary thresholds and a polythetic approach, resulting in thousands of unique symptom combinations for the same diagnosis (Galatzer-Levy & Bryant, 2013). These limitations complicate clinical care and constrain progress in psychiatric nosology and genetics.

To address these issues, approaches such as the Hierarchical Taxonomy of Psychopathology (HiTOP) and the Research Domain Criteria (RDoC) initiative (Cuthbert, 2015; Kotov et al., 2017; Kotov et al., 2021; Kozak & Cuthbert, 2016) have been proposed. HiTOP conceptualizes psychopathology along a continuum from symptoms to broad spectra, culminating in an overarching general psychopathology (*p*) factor. In contrast, RDoC focuses on transdiagnostic domains of functioning to identify putative causal mechanisms across multiple units of analysis. Despite different approaches, HiTOP and RDoC both aim to move beyond traditional diagnostic categories to capture the complexity of psychopathology (Michelini, Palumbo, DeYoung, Latzman, & Kotov, 2021). However, the integration of these frameworks, particularly through genetic investigations, remains largely unexplored.

Genetic research has advanced our understanding of psychopathology dimensions, with most of this work focusing on externalizing (EXT) and internalizing (INT) behaviors. Twin and family studies have shown that both externalizing (e.g., antisocial behavior, substance use) and internalizing (e.g., anxiety, depression) behaviors have shared genetic bases (Hewitt, Silberg, Neale, Eaves, & Erickson, 1992; Silberg et al., 1994). Additionally, twin and family studies (Allegrini et al., 2020; Lahey, Van Hulle, Singh, Waldman, & Rathouz, 2011; Pettersson, Larsson, & Lichtenstein, 2016) and principal component analyses (Allegrini et al., 2020; Selzam, Coleman, Caspi, Moffitt, & Plomin, 2018) have examined genetic factors shared by *both* EXT and INT psychopathology. Genome-wide association studies (GWAS) of childhood behavior problems, encompassing EXT and INT psychopathology, identified two genome-wide significant loci (Neumann et al., 2022; Pappa et al., 2015). These findings align with HiTOP, which posits shared genetic factors within and across psychopathology dimensions, but also highlight the need for larger, more integrative studies to disentangle shared and unique genetic influences across EXT and INT.

Genomic structural equation modeling (gSEM) provides a nuanced understanding of the genetic architecture underlying psychopathology. Studies using gSEM to investigate the factor structure of psychiatric disorders have identified one to four factors that underlie their shared liability (Grotzinger et al., 2022; Grotzinger et al., 2019; Lee et al., 2019). This work has yielded two genome-wide loci for a higher-order *p*-factor that encompasses compulsive, psychotic, internalizing, and neurodevelopmental disorders (Grotzinger et al., 2022), with the researchers noting that the *p*-factor may have low utility for identifying underlying genetic variation. However, because the study included only psychiatric disorders, it did not capture a spectrum of psychopathology consistent with dimensional models like HiTOP. It also included only two EXT (attention-deficit hyperactivity disorder and problematic alcohol use) and two INT (anxiety and major depressive disorder) conditions.

Despite the valuable insights provided by gSEM studies focused specifically on EXT and INT, gaps remain. First, although subclinical traits have been included in studies of EXT (Karlsson Linnér et al., 2019; Karlsson Linnér et al., 2021), their integration in the study of INT has been limited, which risks overlooking genetic contributions across the full range of symptom severity. Second, studies have not typically focused on genetic effects shared across *both* EXT and INT psychopathology. Thus, our understanding of the common genetic factors that drive comorbidities between these two spectra is limited, despite evidence that individuals who experience comorbid EXT and INT problems are uniquely vulnerable to poor economic and social outcomes (Vergunst et al., 2023), criminality (Commisso et al., 2024), and suicide attempts (Commisso et al., 2023).

The current study addresses gaps in our understanding of the genetic architecture of EXT and INT psychopathology. Using gSEM, we tested competing confirmatory factor models based on existing theories of psychopathology to identify the best-fitting structure that captured shared and specific genetic liabilities across clinical and subclinical EXT and INT traits. After identifying the optimal model, we performed GWAS to identify genetic variants that contribute to EXT, INT, and their shared etiology. To further characterize the shared genetic architecture, we applied MiXeR models to estimate polygenicity (i.e., the number of causal variants) and the degree of causal variant overlap between EXT and INT. We also calculated genetic correlations with a wide range of phenotypes to explore the behavioral manifestations of EXT, INT, and EXT+INT genetic liability. These analyses provide insights into the shared and distinct genetic effects that underlie EXT and INT psychopathology and lay the foundation for Part II, a companion article, in which we explore shared and distinct biological mechanisms, including pathways at the levels of genes, cells, and neural circuits.

## Methods

### Summary Statistics

We selected EXT summary statistics based on published studies of individuals genetically similar to Europeans (EUR) (Karlsson Linnér et al., 2019; Karlsson Linnér et al., 2021) and existing theory (Kotov et al., 2017) (Supplementary Table 1). We included the largest GWAS of attention deficit hyperactivity disorder (ADHD; n = 225,534; (Demontis et al., 2023) and four substance use disorders [SUDs i.e., alcohol (AUD; n = 753,248; (Zhou et al., 2023), cannabis (CanUD; n = 886,025; (Levey et al., 2023), opioid (OUD; n = 425,944; (Kember et al., 2022), and tobacco (TUD; n = 495,005; (Toikumo et al., 2023)]. These disorders align with HiTOP’s disinhibited externalizing spectrum (Krueger et al., 2021). We also included subclinical measures reflecting disinhibition and antagonism [age of first sexual intercourse (AgeSex; reverse-coded; n = 317,694), general risk tolerance (Risk; n = 431,126), number of sexual partners (NumSex; n = 370,711; (Karlsson Linnér et al., 2019; Karlsson Linnér et al., 2021), antisocial behavior (ASB; n = 16,400; (Tielbeek et al., 2017), and automobile speeding propensity (n = 404,291; (Karlsson Linnér et al., 2019)].

We selected INT summary statistics that capture HiTOP’s internalizing symptom components, including eating pathology, fear, and distress (Kotov et al., 2017). The studies included the largest available GWAS of INT disorders: (1) anorexia nervosa (AN; n = 72,517; (Watson et al., 2019), (2) major depressive disorder (MDD; n = 1,074,629; (Als et al., 2023), and (3) posttraumatic stress disorder (PTSD; n = 214,408; (Stein et al., 2021). We also included irritability (http://www.nealelab.is/uk-biobank/; n = 345,231), loneliness (n = 490,689; (Abdellaoui et al., 2019), subjective wellbeing (reverse-coded; n = 298,420; (Okbay et al., 2016), miserableness (http://www.nealelab.is/uk-biobank/; n = 355,182), and anxiety (ANX; n = 280,490). To detect variants associated with anxiety disorders (i.e., panic disorder, agoraphobia, generalized anxiety disorder (GAD), specific phobia, and social phobia) and subclinical anxiety (i.e., GAD-2 scores), we combined three GWAS (Levey et al., 2020; Otowa et al., 2016; Purves et al., 2020) via multi-trait analysis of GWAS (Turley et al., 2018).

### Genomic Structural Equation Modeling

We used linkage disequilibrium score regression (LDSC) (Bulik-Sullivan et al., 2015), which estimates the genetic similarity between traits, to calculate genetic correlations (*r*_g_). GenomicSEM is robust to sample overlap, yielding valid results even when overlapping cohorts are included (Grotzinger et al., 2019). Genetic variants, or single nucleotide variants (SNVs), were filtered using reference data from individuals genetically similar to Europeans (EUR HapMap3) (The International HapMap 3 Consortium, 2010). We excluded rare variants (i.e., those present in fewer than 1% of people). LDSC accounts for differences in genetic similarity across and within populations by using linkage disequilibrium (LD) information—patterns of correlation between nearby genetic variants—from the 1000 Genomes Project (The 1000 Genomes Project Consortium, 2015). Traits with an average *r_g_* < 0.20, indicating a small effect size, were excluded from subsequent modeling.

To investigate patterns of genetic overlap, we fit five confirmatory factor analyses (CFAs) based on existing theories of psychopathology (Caspi et al., 2013; Krueger & Markon, 2006; Markon, 2019). Whereas our objective was to test theoretically driven models, we did not perform exploratory factor analyses. First, we evaluated a correlated factors model with two factors representing EXT and INT psychopathology. Next, we tested a three-factor model to explore whether SUDs were distinct from other EXT behaviors. The third model was a bifactor model consisting of a general psychopathology factor and uncorrelated EXT and INT factors. Fourth, we evaluated a higher-order model, which is mathematically equivalent to the two-factor model but represents shared genetic effects between EXT and INT as a second-order factor. To ensure that the model was statistically identified, we constrained the loadings onto the second-order factor to the square root of the genetic correlation between EXT and INT (Loehlin, 1996). Finally, we tested a unidimensional (*p*-factor) model where all traits loaded onto one factor. In all CFA models except the three-factor model (where shared variance is explained by the SUD factor), residual variances for SUDs were allowed to correlate, reflecting shared variance not explained by the broader EXT factor. Model fit was evaluated using chi-square, Akaike information criterion (AIC), comparative fit index (CFI; > 0.90), and standardized root mean squared residual (SRMR; < 0.08) statistics. Factor loadings of ≥ 0.35 were required to ensure that each trait meaningfully contributed to the factor.

### Genome-Wide Association Studies

For GWAS, we excluded rare genetic variants (i.e., those present in <1% of people) and poorly imputed variants (i.e., imputation scores <0.6). Each genetic variant was regressed onto the latent variable(s) using diagonally weighted least squares estimation. To evaluate heterogeneity, we calculated Q_SNV_ values, which indicate whether a variant’s effect is consistent across all traits included on a factor or varies across them. Genetic variants with a Q-statistic p-value < 5x10^-8^ were removed to ensure that only variants with common effects across the spectrum were represented. We identified lead genetic variants using LD clumping in PLINK 1.9 (Purcell et al., 2007), which groups correlated variants based on reference data from the 1000 Genomes Project and identifies the most significant variant in the group.

### Polygenicity and Genetic Overlap

We estimated the genetic complexity of traits using MiXeR, which provides a measure of polygenicity (the number of genetic variants that influence a trait) and discoverability (the average strength of these effects) (Frei et al., 2019; Holland et al., 2020). In MiXeR, causal variants are defined as genetic variants with nonzero additive effects. The method assumes that effects follow a mixture distribution, where a proportion of variants have an effect and others have no effect. MiXeR was also used to identify the proportion of unique and shared causal variants for EXT and INT, regardless of whether their effects are in the same or opposite direction for each trait. Simulation studies have demonstrated that MiXeR is robust to sample overlap (Frei et al., 2019).

### Genetic Correlations

To identify behavioral tendencies that are genetically linked to the spectra, we calculated genetic correlations between EXT, INT, EXT+INT, and 1,437 other traits using LDSC. LDSC accounts for sample overlap to ensure unbiased estimates (Bulik-Sullivan et al., 2015). To account for multiple testing and reduce the risk of false positives, we applied a Benjamini-Hochberg false discovery rate (FDR) correction. We also identified genetic correlations that differed in magnitude for EXT and INT using an FDR correction to determine significance, and determined the proportion of genetic correlations concordant in direction for EXT and INT.

## Results

### Confirmatory Factor Analysis

Of 18 traits examined (Figure 1), two (automobile speeding propensity and anorexia nervosa) were excluded due to weak associations with all others (mean *r*_g_ < 0.20). Using the remaining 16 traits, we tested several CFA models (Figure 2 and Supplementary Figures 1-3). A *p*-factor model did not provide adequate fit (X^2^(98) = 8965.28, AIC = 9041.28, CFI = 0.79, and SRMR = 0.15). Although a bifactor model fit well (X^2^(88) = 3472.02, AIC = 3568.02, CFI = 0.92, and SRMR = 0.07), it led to several weak (< 0.35) and one negative standardized loading. Because of its limited interpretability and several traits that did not adequately load onto the factors, we did not perform GWAS on the bifactor model.

**Figure 1.**
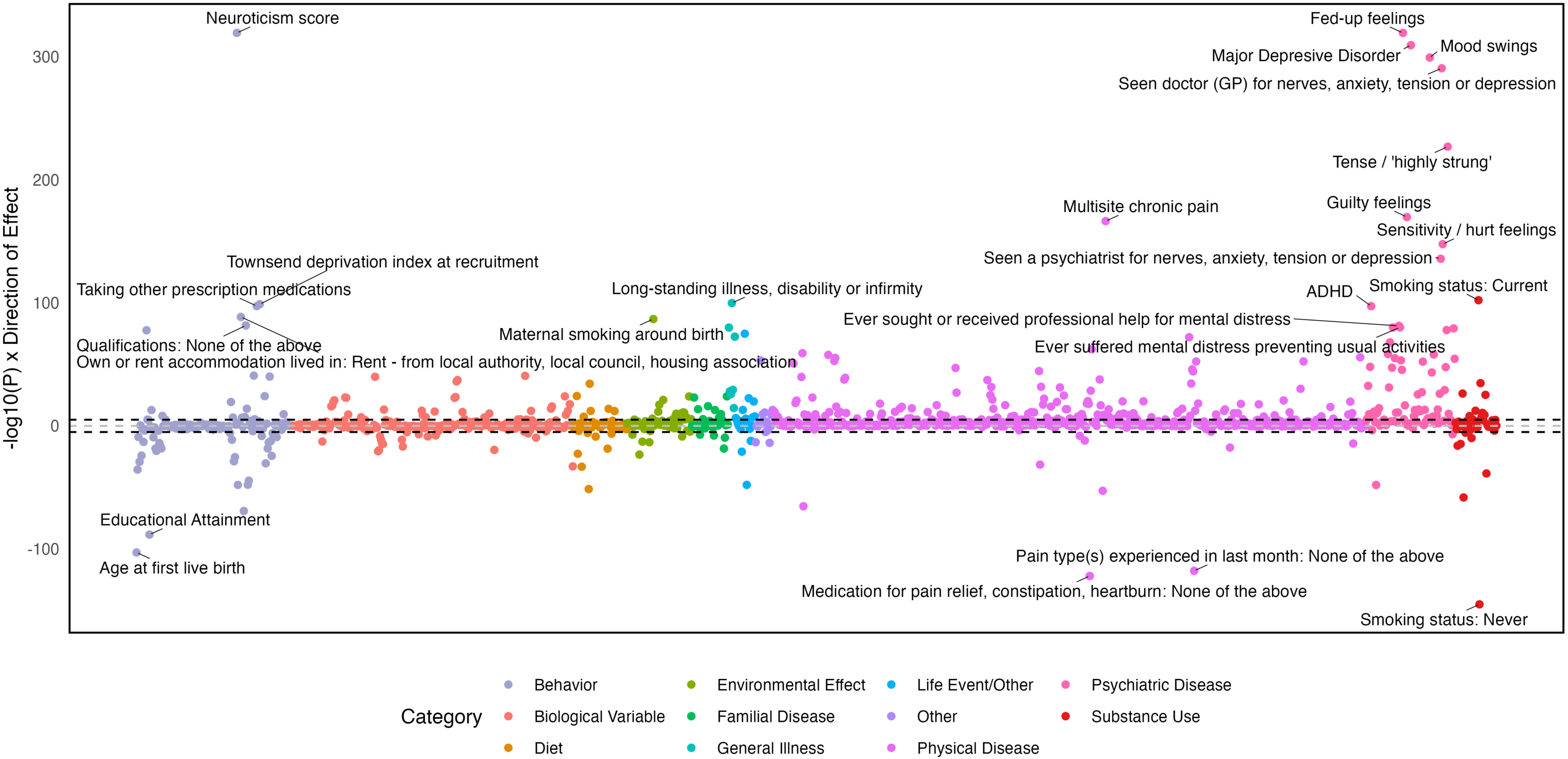
Genetic correlations between the externalizing and internalizing traits. ADHD = attention deficit hyperactivity disorder, AgeSex = age at first sexual intercourse (reverse coded), ASB = antisocial behavior, AUD = alcohol use disorder, CanUD = cannabis use disorder, MDD = major depressive disorder, NumSex = number of sexual partners, OUD = opioid use disorder, PTSD = posttraumatic stress disorder, Risk = general risk tolerance, Speeding = automobile speeding propensity, TUD = tobacco use disorder. Traits are ordered alphabetically. AgeSex and Wellbeing are reverse-coded.

**Figure 2.**
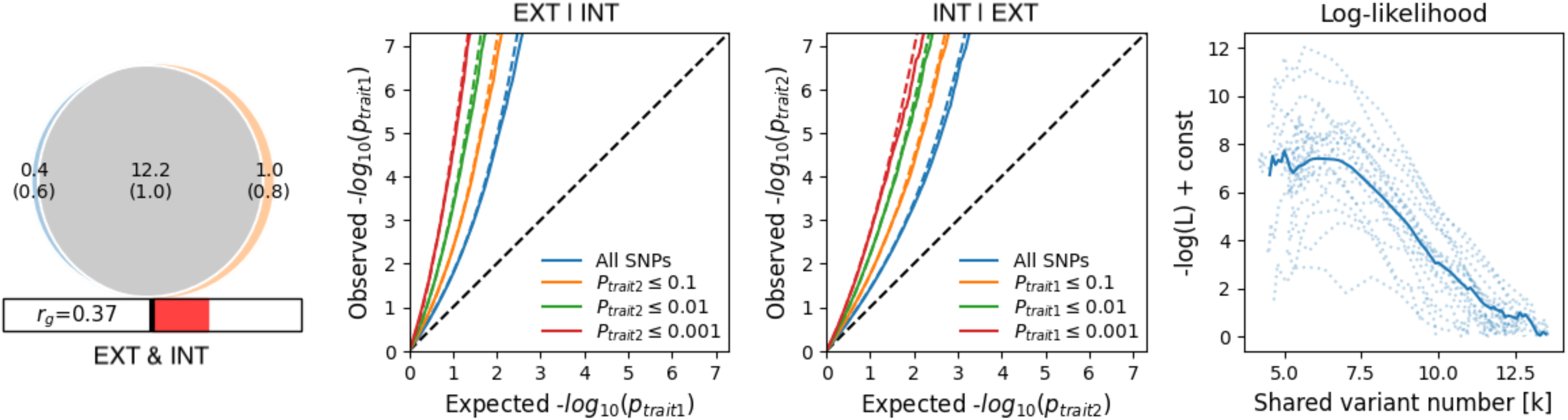
Confirmatory factor analyses of externalizing and internalizing psychopathology. **a)** correlated two-factor model, **b)** higher order factor model. Fit of both: X^2^(97) = 3877.82, AIC = 3955.82, CFI = 0.91, SRMR = 0.09. EXT = externalizing, INT = internalizing, ADHD = attention deficit hyperactivity disorder, AgeSex = age at first sex (reverse coded), NumSex = number of sexual partners, ASB = antisocial behavior, AUD = alcohol use disorder, CanUD = cannabis use disorder, OUD = opioid use disorder, TUD = tobacco use disorder, SWB = subjective wellbeing (reverse coded), PTSD = posttraumatic stress disorder, MDD = major depressive disorder, ANX = anxiety.

Two mathematically equivalent models provided adequate fit and factor loadings: (1) a two-factor correlated model and (2) a higher-order factor model. Fit statistics were X^2^(97) = 3877.82, AIC = 3955.82, CFI = 0.91, and SRMR = 0.09. To ensure identification in the higher-order model, the loadings onto the second-order factor were constrained equal to the square root of the genetic correlation between EXT and INT (Loehlin, 1996), a commonly accepted approach for testing a factor that explains shared variance between two correlated dimensions. Although a three-factor model separating SUDs from other externalizing traits provided reasonable fit (X^2^(101) = 4401.18, AIC = 4471.18, CFI = 0.90, and SRMR = 0.09), it did not improve the fit of the simpler two-factor model and thus did not justify the added complexity.

### Genome-Wide Association Studies

Eighty-three significant SNVs exhibited heterogeneous effects across the EXT spectrum. A plurality was most strongly associated with AgeSex (47.62%), followed by TUD (16.67%; Supplementary Table 2). After removing heterogenous SNVs, the EXT GWAS identified 409 lead SNVs (Supplementary Table 3). Of these, 92 (22.49%) were not identified or located nearby (within ±1000 kb) SNVs identified by the input GWAS, and four were not previously associated with any EXT trait. Three of the novel SNVs were on chromosome 4 (rs1961547, rs9316, and rs7682762), with the fourth on chromosome 22 (rs1473811). These SNVs were previously associated with chronotype, schizophrenia, and social support, among other traits (Supplementary Table 4).

For INT, 41 significant SNVs exhibited heterogeneity, with most (78.05%) showing the strongest associations with MDD (Supplementary Table 5). Removing heterogeneous SNVs left 85 lead SNVs (Supplementary Table 6). Of these, 23 (27.06%) were in loci not significant in the input GWAS, and two were not previously associated with INT (Supplementary Table 7). The novel SNVs were on chromosomes 3 and 4 (rs1381763 and rs4698408).

For EXT+INT, 39 significant SNVs exhibited heterogeneous effects (Supplementary Table 8). Of these, a plurality (41.03%) was most strongly associated with AgeSex, followed by AUD (17.95%). Removing heterogeneous SNVs left 256 lead SNVs (Figure 3 and Supplementary Table 9), 38 of which (14.84%) were not in loci identified by any of the input GWAS, though all were previously associated with an EXT or INT phenotype.

**Figure 3.**
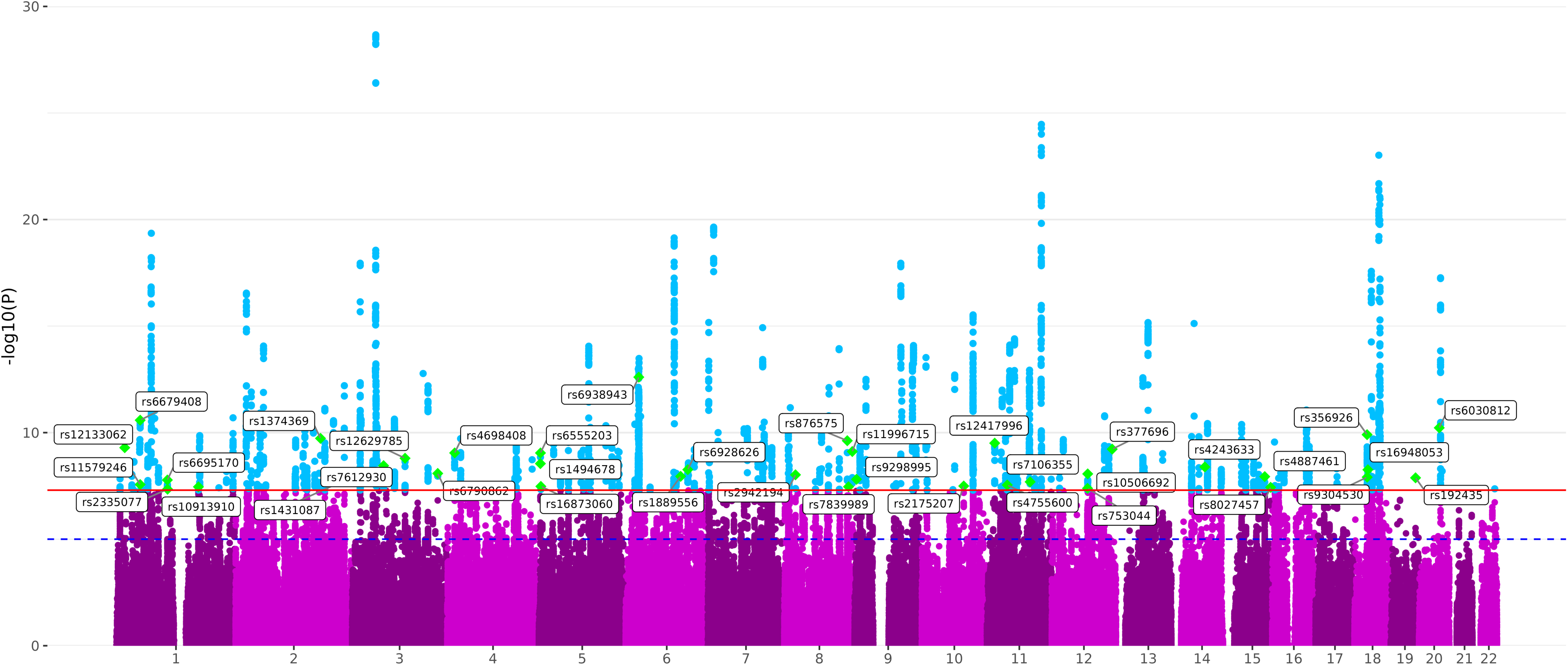
Manhattan plot of the GWAS results for externalizing and internalizing (EXT+INT) liability. Significant single nucleotide variants (SNVs) are highlighted in blue. Green diamonds and annotations denote the lead SNVs in loci not identified in the input GWAS for either of the two spectra.

### Polygenicity and Genetic Overlap

The EXT and INT spectra displayed similar levels of genetic complexity. However, INT had lower discoverability 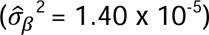 than EXT 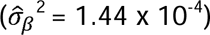, suggesting that INT may require larger samples to detect significant SNVs. Despite an estimated *r_g_* of 0.37, almost all causal variants (96.83% of EXT and 92.42% of INT; Figure 4) overlapped. Of the shared causal variants, 62.92% were estimated to have concordant effect directions.

**Figure 4.**
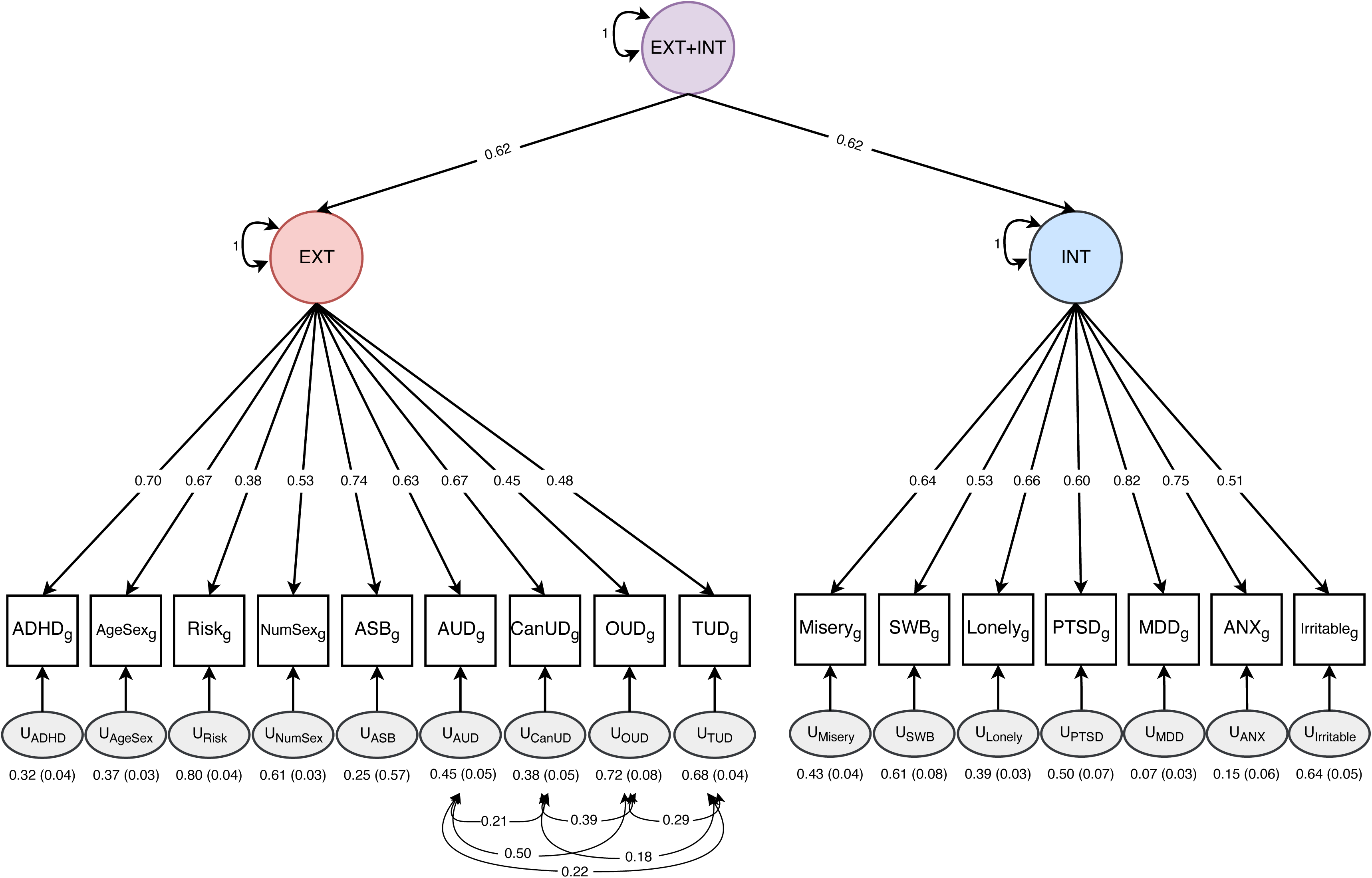
Results of bivariate causal mixture models for externalizing (EXT) and internalizing (INT). The Venn diagram on the left shows the estimated overlap in causal variants for externalizing and internalizing. The next two panels show conditional Q-Q plots of the observed vs. expected -log_10_ p-values in trait 1 as a function of the significance of the association with trait 2 (and vice versa) at the level of p< 0.1, p< 0.01, and p≤ 0.001. The final panel shows the negative log-likelihood as a function of polygenic overlap. The minimum model is represented by the point furthest to the left where the genetic overlap is estimated to be the minimum required to explain the genetic correlation between the two traits. The maximum model is represented as the point furthest on the right represents complete overlap where the least polygenic trait’s causal variants are a subset of the more polygenic trait. The best model (i.e., the one selected by the MiXeR analysis) is the lowest point on the chart.

### Genetic Correlations

There were 413 significant *r_g_* with EXT (Supplementary Table 10 and Supplementary Figure 4), among which tobacco phenotypes were among the strongest (current smoking: *r*_g_ = 0.79, SE = 0.02; ever smoked: *r*_g_ = 0.62, SE = 0.02), as were those with lower socioeconomic status (Townsend deprivation index: *r*_g_ = 0.68, SE = 0.03; financial difficulties: *r*_g_ = 0.58, SE = 0.03) and educational attainment (*r*_g_ = -0.44, SE = 0.02). There were 311 phenotypes genetically correlated with INT (Supplementary Table 11 and Supplementary Figure 5), with the strongest including mood swings (*r*_g_ = 0.90, SE = 0.01) and neuroticism (*r*_g_ = 0.89, SE = 0.01). INT was also genetically correlated with several types of pain (abdominal: *r*_g_ = 0.60, SE = 0.04; facial: *r*_g_ = 0.51, SE = 0.08; chest: *r*_g_ = 0.49, SE = 0.03; and multisite chronic pain: *r*_g_ = 0.49, SE = 0.03). There were 474 significant *r_g_* with EXT+INT, with most being like those of the first-order factors (Supplementary Table 12 and Figure 5). Of the *r_g_*, 17.61% were discordant in direction for EXT and INT, and 168 were of a significantly different magnitude for the two spectra. The *r_g_* with the greatest difference in magnitude were neuroticism score, worrier/anxious feelings, mood swings, fed-up feelings, and worry too long after embarrassment, all of which had stronger positive associations with INT than EXT (Supplementary Figure 6).

**Figure 5.**
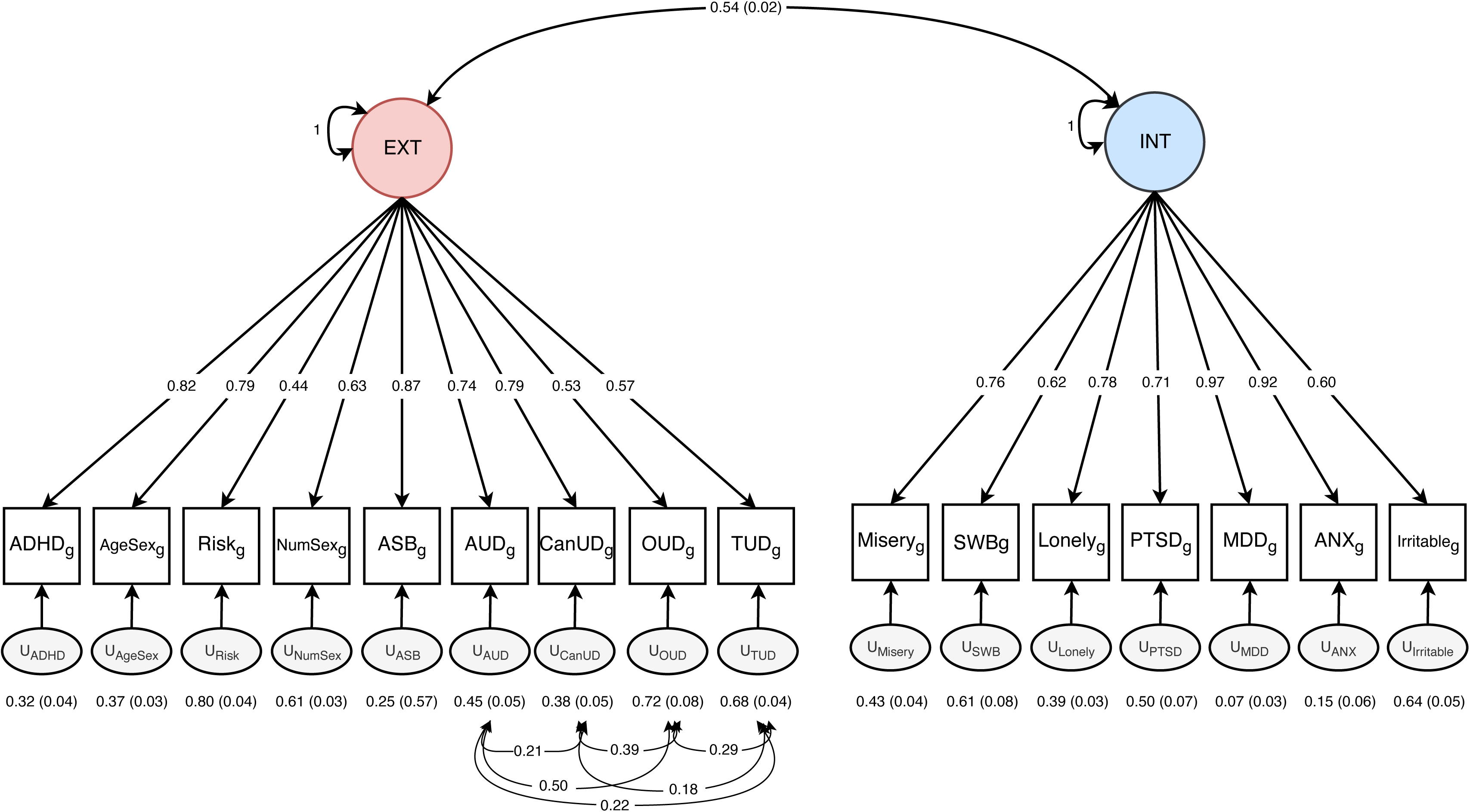
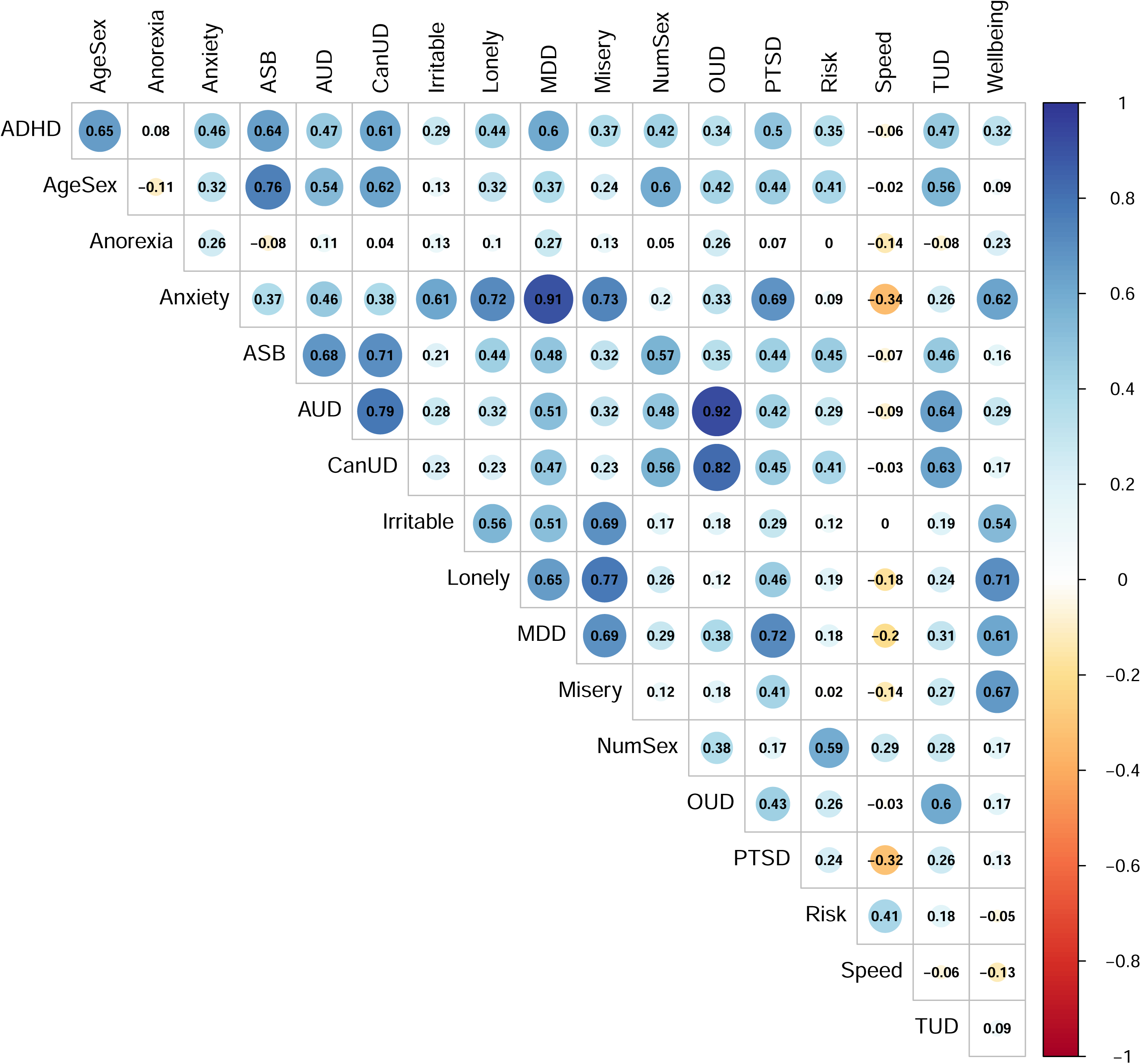
Genetic correlations between the externalizing and internalizing (EXT+INT) factor and publicly available traits. The top 25 associations are annotated.

## Discussion

Using genomic structural equation modeling, we examined the genetic architecture of EXT and INT spectra. Of the factor structures tested, a two-factor correlated model emerged as the best-fitting, with EXT and INT exhibiting a moderate genetic correlation (*r*_g_ = 0.37) that could be modeled as a higher-order EXT+INT factor. GWAS of the latent factors identified 409 independent genomic loci associated with EXT, 85 with INT, and 256 with the higher-order EXT+INT factor. Evaluations of the heterogeneity and polygenicity of each spectrum yielded clear evidence of genetic complexity. Below, we discuss the findings in relation to dimensional models of psychopathology and their implications for biological mechanisms and future research.

### Genetic Insights and Biological Mechanisms

Our findings demonstrate the utility of multivariate approaches for boosting power to detect loci that might otherwise remain undetected when traits are examined in isolation (Grotzinger et al., 2019). By leveraging the shared etiology of commonly co-occurring traits, we identified several novel loci. For both EXT and INT, approximately one-quarter of genetic variants identified were not significant in the GWAS of the individual traits that comprise the spectrum. These results also emphasize the need to study shared liability alongside spectrum-specific effects. For example, 14.84% of loci identified for EXT+INT were captured by neither the EXT or INT factors individually, nor any of the individual trait GWAS. Thus, there is added value in modeling higher-order constructs to capture biological mechanisms that transcend individual traits and spectra. This higher-order factor may represent “genetic convergence,” whereby overlapping genetic influences coalesce to drive the co-occurrence of psychopathology across spectra (Waszczuk et al., 2020).

### Spectrum-Level Heterogeneity and Genetic Complexity

We identified differences in the degree of genetic variability for EXT, INT, and their shared liability (EXT+INT). EXT exhibited the most heterogeneity, with 83 genetic variants showing varying effects across the EXT spectrum. INT, in contrast, showed less heterogeneity, with just 41 genetic variants that had differing effects across INT traits. The higher-order EXT+INT factor showed the least heterogeneity, with 39 genetic variants having differing effects across the spectra. These findings offer insights into two possible frameworks for understanding heterogeneity in hierarchical models of psychopathology (Markon, 2019; Waszczuk et al., 2020).

The first framework is a “top-up” approach, in which the higher-order EXT+INT factor captures and accumulates the heterogeneity present within the two lower-order spectra (Markon, 2019). Given this, we would expect a greater degree of heterogeneity for EXT+INT than the first-order spectra. In contrast, the second framework posits a “top-down” model, whereby the EXT+INT factor reflects a more stable and cohesive genetic architecture than either first-order factor (Markon, 2019). Heterogeneity, in this case, would be more pronounced at lower levels of the hierarchy. Our findings align more closely with this second perspective. Less heterogeneity for EXT+INT than EXT and INT individually suggests that the higher-order spectrum consolidates overlapping genetic influences into a more cohesive signal, challenging the assumption that broader spectra necessarily amplify complexity.

Insights from MiXeR reinforce the interpretation that the EXT+INT factor reflects a cohesive and stable genetic architecture. Despite a modest genetic correlation of 0.37 between EXT and INT, MiXeR estimated that nearly all causal genetic variants that influence EXT (96.83%) and INT (92.42%) were shared across the two spectra. Although many of these variants were estimated to have opposite directions of effect (37.08%), these findings underscore the potential value of higher-order constructs for reducing trait-specific noise, offering a clearer lens through which to understand the genetic etiology of psychopathology.

### Genetic Correlations and Phenotypic Patterns

Genetic correlations of the two spectra revealed distinct but overlapping phenotypes, with discordant directions for 17.61% of *r*_g_ and significant differences in magnitude across 168 traits. EXT was predominantly genetically correlated with substance use and socioeconomic status, while most genetic correlations with INT were with anxiety- and mood-related traits. Despite this divergence, the top *r*_g_ for the two spectra overlapped somewhat, particularly for pain-related phenotypes. EXT+INT was also genetically correlated with pain-related phenotypes, substance use, and mood. Although these patterns support differentiating between the two spectra, they also highlight areas of overlap where common genetic factors exhibit pleiotropy with other behavioral manifestations.

### Implications for Dimensional Models of Psychopathology

Our findings offer important insights for dimensional models of psychopathology like HiTOP and RDoC by supporting and refining their conceptual frameworks. HiTOP emphasizes a hierarchical organization of symptoms and spectra, proposing that broader dimensions like EXT+INT capture shared liability across lower-order spectra (Kotov et al., 2017). Although this aligns with our observation that the EXT+INT factor accounts for shared liability between the EXT and INT spectra, its lower heterogeneity challenges the expectation that heterogeneity accumulates in broader psychopathology spectra. Instead, higher-order psychopathology spectra may reflect more cohesive constructs than previously assumed, necessitating a reevaluation of how hierarchical models conceptualize heterogeneity across levels. Alternatively, the reduced heterogeneity may stem from the modeling approach. Future research should explore whether this pattern is specific to EXT+INT or generalizes to other higher-order psychopathology spectra.

The mechanistic focus of RDoC provides a complementary lens for interpreting our findings. By examining genetic correlations and associated genetic variants for the spectra, we connect HiTOP’s structural dimensions with RDoC’s emphasis on genes and behavior as two units of analysis. The EXT and INT spectra align with distinct RDoC domains. EXT primarily maps onto the positive valence and cognitive systems, as evidenced by genetic correlations with substance use and risky decision making, which reflect processes related to reward sensitivity and impulsivity. In contrast, INT aligns strongly with the negative valence systems and arousal/regulatory systems, as evidenced by genetic correlations with mood swings, neuroticism, and pain, which are linked to threat sensitivity and stress reactivity. Framing higher-order dimensions like EXT+INT as bridges between these distinct RDoC domains begins to unify structural and mechanistic approaches to psychopathology, a key focus of Part II of this pair of companion articles.

## Limitations

A key limitation of this study is the inclusion of only European-ancestry individuals. Although GWAS of EXT- and INT-related *disorders* are available for some non-European ancestry groups, those of more precise, *non-disorder* psychiatric phenotypes are limited. Nonetheless, some research suggests a similar factor structure exists in African-ancestry individuals. At the disorder level, a gSEM of African-ancestry individuals identified substance use and psychiatric disorder factors that roughly aligned with EXT and INT (Khan et al., 2024). Additionally, the analyses showed that a higher-order factor accounted for genetic variance shared by substance use and psychiatric disorders, which to a degree corresponds to our EXT+INT factor. With the growth of diverse biobanks (Bianchi et al., 2024) and deep phenotyping (Sanchez-Roige & Palmer, 2020), direct replication in other ancestries should soon be possible and given high priority.

Another limitation is the study’s focus on EXT and INT, which does not fully capture psychopathology. Future research could incorporate a wider range of traits, including those related to thought or personality disorders, to provide a more comprehensive understanding of the genetic architecture underlying psychopathology. Finally, most existing GWAS are cross-sectional and detailed demographic information is not always reported due to the meta-analytic nature of these studies. For developmental disorders such as ADHD and conduct disorder, cross-sectional approaches in which the ages of individuals represented vary may obscure genetic insights.

## Conclusions

Our findings highlight the value of using genetic data to refine and evaluate dimensional models of psychopathology. By modeling EXT, INT, and their shared liability (EXT+INT), we identified a more cohesive genetic etiology operating at the higher-order level, challenging the assumption that broader spectra necessarily increase heterogeneity. Instead, the EXT+INT factor consolidated overlapping genetic influences, offering a clearer framework for understanding comorbidity. These findings can inform models like HiTOP and RDoC, emphasizing the need to balance shared liability with spectrum-specific effects. Future research, including the companion Part II article, will be crucial for identifying the specific and shared biological pathways that underlie these spectra. The effort to link dimensional models to biological mechanisms will serve to bridge structural (i.e., HiTOP) and mechanistic (i.e., RDoC) approaches to psychopathology, ultimately enhancing their clinical and theoretical utility.

## Financial Support

This work was supported by the Veterans Integrated Service Network 4 Mental Illness Research, Education and Clinical Center and by Department of Veterans Affairs grants I01 BX004820 to H.R.K., National Institute on Alcohol Abuse and Alcoholism grant AA028292 to R.L.K, and KNAW (Royal Netherlands Academy of Arts and Sciences) Academy Professor Award (PAH/6635) to D.I.B. The funders had no role in study design, data collection or analysis, decision to publish, or preparation of the manuscript.

## Competing Interests

Dr. Kranzler is a member of advisory boards for Altimmune, Clearmind Medicine, Dicerna Pharmaceuticals, Enthion Pharmaceuticals, Lilly Pharmaceuticals, and Sophrosyne Pharmaceuticals; a consultant to Altimmune and Sobrera Pharmaceuticals; the recipient of research funding and medication supplies for an investigator-initiated study from Alkermes; and a member of the American Society of Clinical Psychopharmacology’s Alcohol Clinical Trials Initiative, which was supported in the last three years by Alkermes, Dicerna, Ethypharm, Lundbeck, Mitsubishi, Otsuka, and Pear Therapeutics. Drs. Kranzler and Gelernter hold U.S. patent 10,900,082 titled: “Genotype-guided dosing of opioid agonists,” issued 26 January 2021. The other authors have no disclosures to make.

## Data Availability

All data produced in the present study are available upon reasonable request to the authors. Upon acceptance in a peer-reviewed journal, summary statistics produced will be made publicly available.

## Notes

### Author Declarations

GWAS Summary Statistics used for the present study can be accessed at the following locations: UK Biobank (http://www.nealelab.is/uk-biobank/), dbGaP accession phs001672 for Million Veteran Program (https://www.ncbi.nlm.nih.gov/projects/gap/cgi-bin/study.cgi?study_id=phs001672), iPSYCH (https://ipsych.dk/en/research/downloads/), Psychiatric Genetics Consortium (https://pgc.unc.edu/for-researchers/download-results/), the Social Science Genetic Association Consortium (https://thessgac.com/), GWAS catalog (https://www.ebi.ac.uk/gwas/publications/28979981; https://www.ebi.ac.uk/gwas/publications/33532862), Gelernter Lab website (https://medicine.yale.edu/lab/gelernter/stats/), Global Biobank Meta-analysis Initiative (https://www.globalbiobankmeta.org/resources), and the Diabetes Genetics Replication and Meta-Analysis Consortium (https://diagram-consortium.org/downloads.html).

### Summary of Updates

Original manuscript has been divided into two companion manuscripts.

## References

Abdellaoui, A., Sanchez-Roige, S., Sealock, J., Treur, J. L., Dennis, J., Fontanillas, P., . . . Boomsma, D. I. (2019). Phenome-wide investigation of health outcomes associated with genetic predisposition to loneliness. Human Molecular Genetics, 28(22), 3853–3865. doi:10.1093/hmg/ddz219

Allegrini, A. G., Cheesman, R., Rimfeld, K., Selzam, S., Pingault, J.-B., Eley, T. C., & Plomin, R. (2020). The p factor: genetic analyses support a general dimension of psychopathology in childhood and adolescence. Journal of Child Psychology and Psychiatry, 61(1), 30–39. doi:10.1111/jcpp.13113

Als, T. D., Kurki, M. I., Grove, J., Voloudakis, G., Therrien, K., Tasanko, E., . . . Børglum, A. D. (2023). Depression pathophysiology, risk prediction of recurrence and comorbid psychiatric disorders using genome-wide analyses. Nature Medicine, 29(7), 1832–1844. doi:10.1038/s41591-023-02352-1

Bianchi, D. W., Brennan, P. F., Chiang, M. F., Criswell, L. A., D’Souza, R. N., Gibbons, G. H., . . . Denny, J. C. (2024). The All of Us Research Program is an opportunity to enhance the diversity of US biomedical research. Nature Medicine, 30(2), 330–333. doi:10.1038/s41591-023-02744-3

Bulik-Sullivan, B. K., Loh, P.-R., Finucane, H. K., Ripke, S., Yang, J., Patterson, N., . . . Schizophrenia Working Group of the Psychiatric Genomics, C. (2015). LD Score regression distinguishes confounding from polygenicity in genome-wide association studies. Nature Genetics, 47(3), 291-295. doi:10.1038/ng.3211

Caspi, A., Houts, R. M., Belsky, D. W., Goldman-Mellor, S. J., Harrington, H., Israel, S., . . . Moffitt, T. E. (2013). The p factor: One general psychopathology factor in the structure of psychiatric disorders? Clinical Psychological Science, 2(2), 119–137. doi:10.1177/2167702613497473

Commisso, M., Geoffroy, M.-C., Temcheff, C., Scardera, S., Vergunst, F., Côté, S. M., . . . Orri, M. (2024). Association of childhood externalizing, internalizing, comorbid problems with criminal convictions by early adulthood. Journal of Psychiatric Research, 172, 9–15. doi:10.1016/j.jpsychires.2024.01.039

Commisso, M., Temcheff, C., Orri, M., Poirier, M., Lau, M., Côté, S., . . . Geoffroy, M.-C. (2023). Childhood externalizing, internalizing and comorbid problems: distinguishing young adults who think about suicide from those who attempt suicide. Psychological Medicine, 53(3), 1030–1037. doi:10.1017/S0033291721002464

Cuthbert, B. N. (2015). Research Domain Criteria: Toward future psychiatric nosologies. Dialogues in Clinical Neuroscience, 17(1), 89–97. doi:10.31887/DCNS.2015.17.1/bcuthbert

Demontis, D., Walters, G. B., Athanasiadis, G., Walters, R., Therrien, K., Nielsen, T. T., . . . Børglum, A. D. (2023). Genome-wide analyses of ADHD identify 27 risk loci, refine the genetic architecture and implicate several cognitive domains. Nature Genetics, 55(2), 198–208. doi:10.1038/s41588-022-01285-8

Frei, O., Holland, D., Smeland, O. B., Shadrin, A. A., Fan, C. C., Maeland, S., . . . Dale, A. M. (2019). Bivariate causal mixture model quantifies polygenic overlap between complex traits beyond genetic correlation. Nature Communications, 10(1), 2417. doi:10.1038/s41467-019-10310-0

Galatzer-Levy, I. R., & Bryant, R. A. (2013). 636,120 ways to have posttraumatic stress disorder. Perspectives on Psychological Science, 8(6), 651–662. doi:10.1177/1745691613504115

Grotzinger, A. D., Mallard, T. T., Akingbuwa, W. A., Ip, H. F., Adams, M. J., Lewis, C. M., . . . Schizophrenia Working Group of the Psychiatric Genetics, C. (2022). Genetic architecture of 11 major psychiatric disorders at biobehavioral, functional genomic and molecular genetic levels of analysis. Nature Genetics, 54(5), 548-559. doi:10.1038/s41588-022-01057-4

Grotzinger, A. D., Rhemtulla, M., de Vlaming, R., Ritchie, S. J., Mallard, T. T., Hill, W. D., . . . Tucker-Drob, E. M. (2019). Genomic structural equation modelling provides insights into the multivariate genetic architecture of complex traits. Nature Human Behaviour, 3(5), 513–525. doi:10.1038/s41562-019-0566-x

Hewitt, J. K., Silberg, J. L., Neale, M. C., Eaves, L. J., & Erickson, M. (1992). The analysis of parental ratings of children’s behavior using LISREL. Behavior Genetics, 22(3), 293–317. doi:10.1007/BF01066663

Holland, D., Frei, O., Desikan, R., Fan, C.-C., Shadrin, A. A., Smeland, O. B., . . . Dale, A. M. (2020). Beyond SNP heritability: Polygenicity and discoverability of phenotypes estimated with a univariate Gaussian mixture model. PLOS Genetics, 16(5), e1008612. doi:10.1371/journal.pgen.1008612

Karlsson Linnér, R., Biroli, P., Kong, E., Meddens, S. F. W., Wedow, R., Fontana, M. A., . . . Beauchamp, J. P. (2019). Genome-wide association analyses of risk tolerance and risky behaviors in over 1 million individuals identify hundreds of loci and shared genetic influences. Nature Genetics, 51(2), 245–257. doi:10.1038/s41588-018-0309-3

Karlsson Linnér, R., Mallard, T. T., Barr, P. B., Sanchez-Roige, S., Madole, J. W., Driver, M. N., . . . Collaborators, C. (2021). Multivariate analysis of 1.5 million people identifies genetic associations with traits related to self-regulation and addiction. Nature Neuroscience, 24(10), 1367–1376. doi:10.1038/s41593-021-00908-3

Kember, R. L., Vickers-Smith, R., Xu, H., Toikumo, S., Niarchou, M., Zhou, H., . . . Million Veteran, P. (2022). Cross-ancestry meta-analysis of opioid use disorder uncovers novel loci with predominant effects in brain regions associated with addiction. Nature Neuroscience, 25(10), 1279–1287. doi:10.1038/s41593-022-01160-z

Kessler, R. C., Chiu, W. T., Demler, O., & Walters, E. E. (2005). Prevalence, severity, and comorbidity of 12-month DSM-IV disorders in the National Comorbidity Survey replication. Archives of General Psychiatry, 62(6), 617–627. doi:10.1001/archpsyc.62.6.617

Kessler, R. C., McGonagle, K. A., Zhao, S., Nelson, C. B., Hughes, M., Eshleman, S., . . . Kendler, K. S. (1994). Lifetime and 12-month prevalence of DSM-III-R psychiatric disorders in the United States: Results from the National Comorbidity Survey. Archives of General Psychiatry, 51(1), 8–19. doi:10.1001/archpsyc.1994.03950010008002

Khan, Y., Davis, C. N., Jinwala, Z., Feuer, K. L., Toikumo, S., Hartwell, E. E., . . . Kember, R. L. (2024). Combining transdiagnostic and disorder-level GWAS enhances precision of psychiatric genetic risk profiles in a multi-ancestry sample. medRxiv. doi:10.1101/2024.05.09.24307111

Kotov, R., Krueger, R. F., Watson, D., Achenbach, T. M., Althoff, R. R., Bagby, R. M., . . . Zimmerman, M. (2017). The Hierarchical Taxonomy of Psychopathology (HiTOP): A dimensional alternative to traditional nosologies. Journal of Abnormal Psychology, 126(4), 454–477. doi:10.1037/abn0000258

Kotov, R., Krueger, R. F., Watson, D., Cicero, D. C., Conway, C. C., DeYoung, C. G., . . . Wright, A. G. C. (2021). The Hierarchical Taxonomy of Psychopathology (HiTOP): A quantitative nosology based on consensus of evidence. Annual Review of Clinical Psychology, 17(1), 83–108. doi:10.1146/annurev-clinpsy-081219-093304

Kozak, M. J., & Cuthbert, B. N. (2016). The NIMH Research Domain Criteria Initiative: Background, issues, and pragmatics. Psychophysiology, 53(3), 286–297. doi:10.1111/psyp.12518

Krueger, R. F., Hobbs, K. A., Conway, C. C., Dick, D. M., Dretsch, M. N., Eaton, N. R., . . . HiTOP Utility Workgroup (2021). Validity and utility of Hierarchical Taxonomy of Psychopathology (HiTOP): II. Externalizing superspectrum. World Psychiatry, 20(2), 171–193. doi:10.1002/wps.20844

Krueger, R. F., & Markon, K. E. (2006). Reinterpreting comorbidity: A model-based approach to understanding and classifying psychopathology. Annual Review of Clinical Psychology, 2(Volume 2, 2006), 111-133. doi:10.1146/annurev.clinpsy.2.022305.095213

Lahey, B. B., Van Hulle, C. A., Singh, A. L., Waldman, I. D., & Rathouz, P. J. (2011). Higher-order genetic and environmental structure of prevalent forms of child and adolescent psychopathology. Archives of General Psychiatry, 68(2), 181–189. doi:10.1001/archgenpsychiatry.2010.192

Lee, P. H., Anttila, V., Won, H., Feng, Y.-C. A., Rosenthal, J., Zhu, Z., . . . Smoller, J. W. (2019). Genomic relationships, novel loci, and pleiotropic mechanisms across eight psychiatric disorders. Cell, 179(7), 1469–1482.e1411. doi:10.1016/j.cell.2019.11.020

Lee, P. H., Feng, Y.-C. A., & Smoller, J. W. (2021). Pleiotropy and cross-disorder genetics among psychiatric disorders. Biological Psychiatry, 89(1), 20–31. doi:10.1016/j.biopsych.2020.09.026

Levey, D. F., Galimberti, M., Deak, J. D., Wendt, F. R., Bhattacharya, A., Koller, D., . . . Veterans Affairs Million Veteran, P. (2023). Multi-ancestry genome-wide association study of cannabis use disorder yields insight into disease biology and public health implications. Nature Genetics, 55(12), 2094-2103. doi:10.1038/s41588-023-01563-z

Levey, D. F., Gelernter, J., Polimanti, R., Zhou, H., Cheng, Z., Aslan, M., . . . Stein, M. B. (2020). Reproducible genetic risk loci for anxiety: Results from ∼200,000 participants in the Million Veteran Program. American Journal of Psychiatry, 177(3), 223–232. doi:10.1176/appi.ajp.2019.19030256

Loehlin, J. C. (1996). The Cholesky approach: A cautionary note. Behavior Genetics, 26(1), 65–69. doi:10.1007/BF02361160

Markon, K. E. (2019). Bifactor and hierarchical models: Specification, inference, and interpretation. Annual Review of Clinical Psychology, 15(Volume 15, 2019), 51-69. doi:10.1146/annurev-clinpsy-050718-095522

Michelini, G., Palumbo, I. M., DeYoung, C. G., Latzman, R. D., & Kotov, R. (2021). Linking RDoC and HiTOP: A new interface for advancing psychiatric nosology and neuroscience. Clinical Psychology Review, 86, 102025. doi:10.1016/j.cpr.2021.102025

Neumann, A., Nolte, I. M., Pappa, I., Ahluwalia, T. S., Pettersson, E., Rodriguez, A., . . . Tiemeier, H. (2022). A genome-wide association study of total child psychiatric problems scores. PLoS One, 17(8), e0273116. doi:10.1371/journal.pone.0273116

Okbay, A., Baselmans, B. M. L., De Neve, J.-E., Turley, P., Nivard, M. G., Fontana, M. A., . . . LifeLines Cohort, S. (2016). Genetic variants associated with subjective well-being, depressive symptoms, and neuroticism identified through genome-wide analyses. Nature Genetics, 48(6), 624–633. doi:10.1038/ng.3552

Otowa, T., Hek, K., Lee, M., Byrne, E. M., Mirza, S. S., Nivard, M. G., . . . Hettema, J. M. (2016). Meta-analysis of genome-wide association studies of anxiety disorders. Molecular Psychiatry, 21(10), 1391–1399. doi:10.1038/mp.2015.197

Pappa, I., Fedko, I. O., Mileva-Seitz, V. R., Hottenga, J. J., Bakermans-Kranenburg, M. J., Bartels, M., . . . Boomsma, D. I. (2015). Single nucleotide polymorphism heritability of behavior problems in childhood: Genome-wide complex trait analysis. Journal of the American Academy of Child and Adolescent Psychiatry, 54(9), 737–744. doi:10.1016/j.jaac.2015.06.004

Pettersson, E., Larsson, H., & Lichtenstein, P. (2016). Common psychiatric disorders share the same genetic origin: a multivariate sibling study of the Swedish population. Molecular Psychiatry, 21(5), 717–721. doi:10.1038/mp.2015.116

Purcell, S., Neale, B., Todd-Brown, K., Thomas, L., Ferreira, M. A. R., Bender, D., . . . Sham, P. C. (2007). PLINK: a tool set for whole-genome association and population-based linkage analyses. American Journal of Human Genetics, 81(3), 559–575. doi:10.1086/519795

Purves, K. L., Coleman, J. R. I., Meier, S. M., Rayner, C., Davis, K. A. S., Cheesman, R., . . . Eley, T. C. (2020). A major role for common genetic variation in anxiety disorders. Molecular Psychiatry, 25(12), 3292–3303. doi:10.1038/s41380-019-0559-1

Sanchez-Roige, S., & Palmer, A. A. (2020). Emerging phenotyping strategies will advance our understanding of psychiatric genetics. Nature Neuroscience, 23(4), 475–480. doi:10.1038/s41593-020-0609-7

Selzam, S., Coleman, J. R. I., Caspi, A., Moffitt, T. E., & Plomin, R. (2018). A polygenic p factor for major psychiatric disorders. Translational Psychiatry, 8(1), 205. doi:10.1038/s41398-018-0217-4

Silberg, J. L., Erickson, M. T., Meyer, J. M., Eaves, L. J., Rutter, M. L., & Hewitt, J. K. (1994). The application of structural equation modeling to maternal ratings of twins’ behavioral and emotional problems. Journal of Consulting and Clinical Psychology, 62(3), 510–521.

Stein, M. B., Levey, D. F., Cheng, Z., Wendt, F. R., Harrington, K., Pathak, G. A., . . . Program, V. A. M. V. (2021). Genome-wide association analyses of post-traumatic stress disorder and its symptom subdomains in the Million Veteran Program. Nature Genetics, 53(2), 174–184. doi:10.1038/s41588-020-00767-x

The 1000 Genomes Project Consortium. (2015). A global reference for human genetic variation. Nature, 526(7571), 68-74. doi:10.1038/nature15393

The International HapMap 3 Consortium. (2010). Integrating common and rare genetic variation in diverse human populations. Nature, 467(7311), 52-58. doi:10.1038/nature09298

Tielbeek, J. J., Johansson, A., Polderman, T. J. C., Rautiainen, M.-R., Jansen, P., Taylor, M., . . . collaborators, f. t. B. A. B. C. (2017). Genome-wide association studies of a broad spectrum of antisocial behavior. JAMA Psychiatry, 74(12), 1242-1250. doi:10.1001/jamapsychiatry.2017.3069

Toikumo, S., Jennings, M. V., Pham, B. K., Lee, H., Mallard, T. T., Bianchi, S. B., . . . Sanchez-Roige, S. (2023). Multi-ancestry meta-analysis of tobacco use disorder prioritizes novel candidate risk genes and reveals associations with numerous health outcomes. medRxiv. doi:10.1101/2023.03.27.23287713

Turley, P., Walters, R. K., Maghzian, O., Okbay, A., Lee, J. J., Fontana, M. A., . . . Social Science Genetic Association, C. (2018). Multi-trait analysis of genome-wide association summary statistics using MTAG. *Nature Genetics*, *50*(2), 229-237. doi:10.1038/s41588-017-0009-4

Vergunst, F., Commisso, M., Geoffroy, M.-C., Temcheff, C., Poirier, M., Park, J., . . . Orri, M. (2023). Association of childhood externalizing, internalizing, and comorbid symptoms with long-term economic and social outcomes. JAMA Network Open, 6(1), e2249568–e2249568. doi:10.1001/jamanetworkopen.2022.49568

Waszczuk, M. A., Eaton, N. R., Krueger, R. F., Shackman, A. J., Waldman, I. D., Zald, D. H., . . . Kotov, R. (2020). Redefining phenotypes to advance psychiatric genetics: Implications from hierarchical taxonomy of psychopathology. Journal of Abnormal Psychology, 129(2), 143–161. doi:10.1037/abn0000486

Watson, H. J., Yilmaz, Z., Thornton, L. M., Hübel, C., Coleman, J. R. I., Gaspar, H. A., . . . Eating Disorders Working Group of the Psychiatric Genomics Consortium. (2019). Genome-wide association study identifies eight risk loci and implicates metabo-psychiatric origins for anorexia nervosa. Nature Genetics, 51(8), 1207-1214. doi:10.1038/s41588-019-0439-2

Zhou, H., Kember, R. L., Deak, J. D., Xu, H., Toikumo, S., Yuan, K., . . . Million Veteran Program. (2023). Multi-ancestry study of the genetics of problematic alcohol use in over 1 million individuals. Nature Medicine. doi:10.1038/s41591-023-02653-5

