## Supplementary Figures for "Integrating HiTOP and RDoC Frameworks Part I: Genetic Architecture of Externalizing and Internalizing Psychopathology"

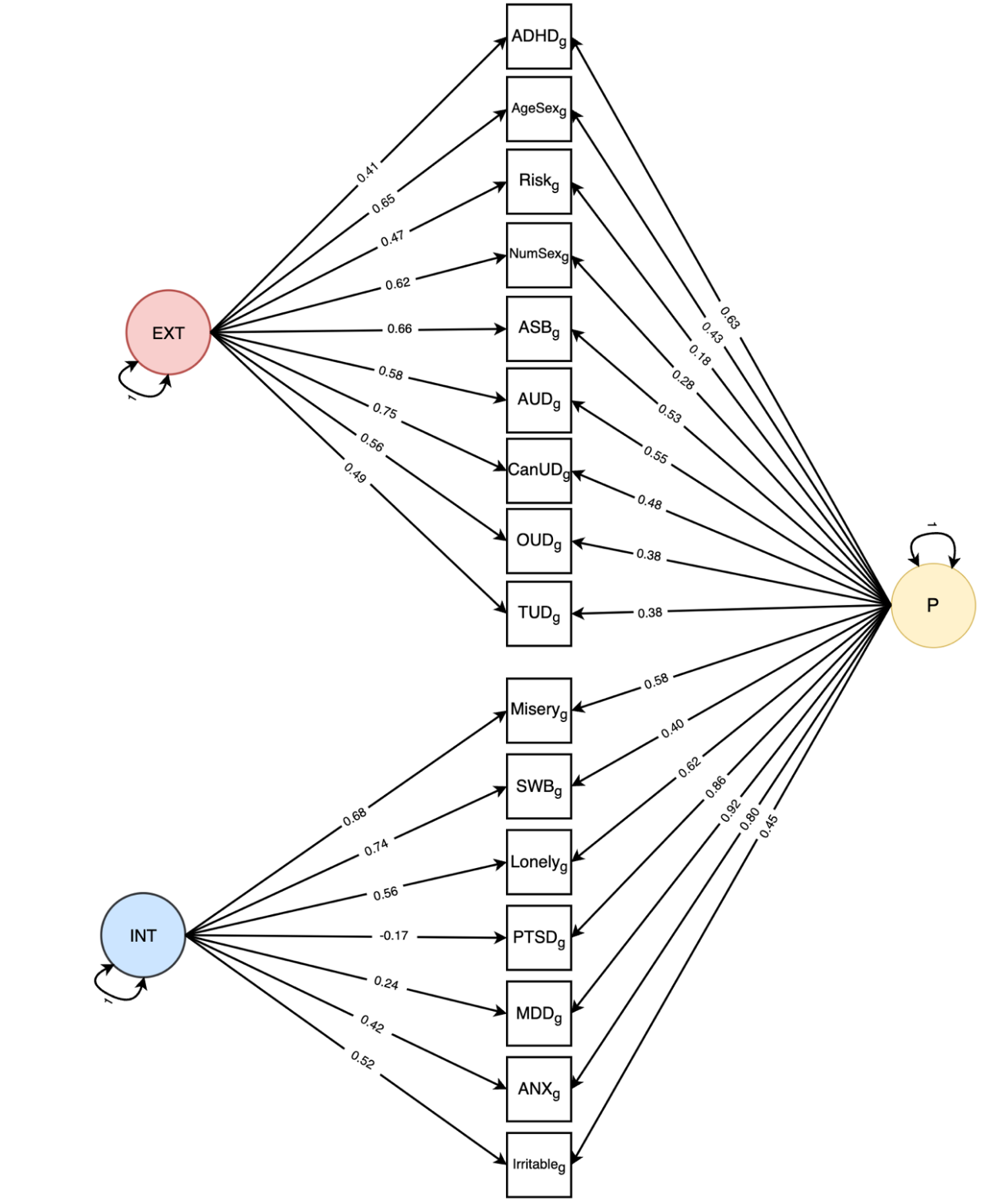

### **Supplementary Figure 1. Bifactor confirmatory factor model.**

Model fit: $\chi$^2^(88) = 3472.02, AIC = 3568.02, CFI = 0.92, and SRMR = 0.07. EXT = externalizing, INT = internalizing, P = general psychopathology, ADHD = attention deficit hyperactivity disorder, AgeSex = age at first sexual intercourse (reverse-coded), NumSex = number of sexual partners, ASB = antisocial behavior, AUD = alcohol use disorder, CanUD = cannabis use disorder, OUD = opioid use disorder, TUD = tobacco use disorder, SWB = subjective wellbeing (reverse-coded), PTSD = posttraumatic stress disorder, MDD = major depressive disorder, ANX = anxiety.

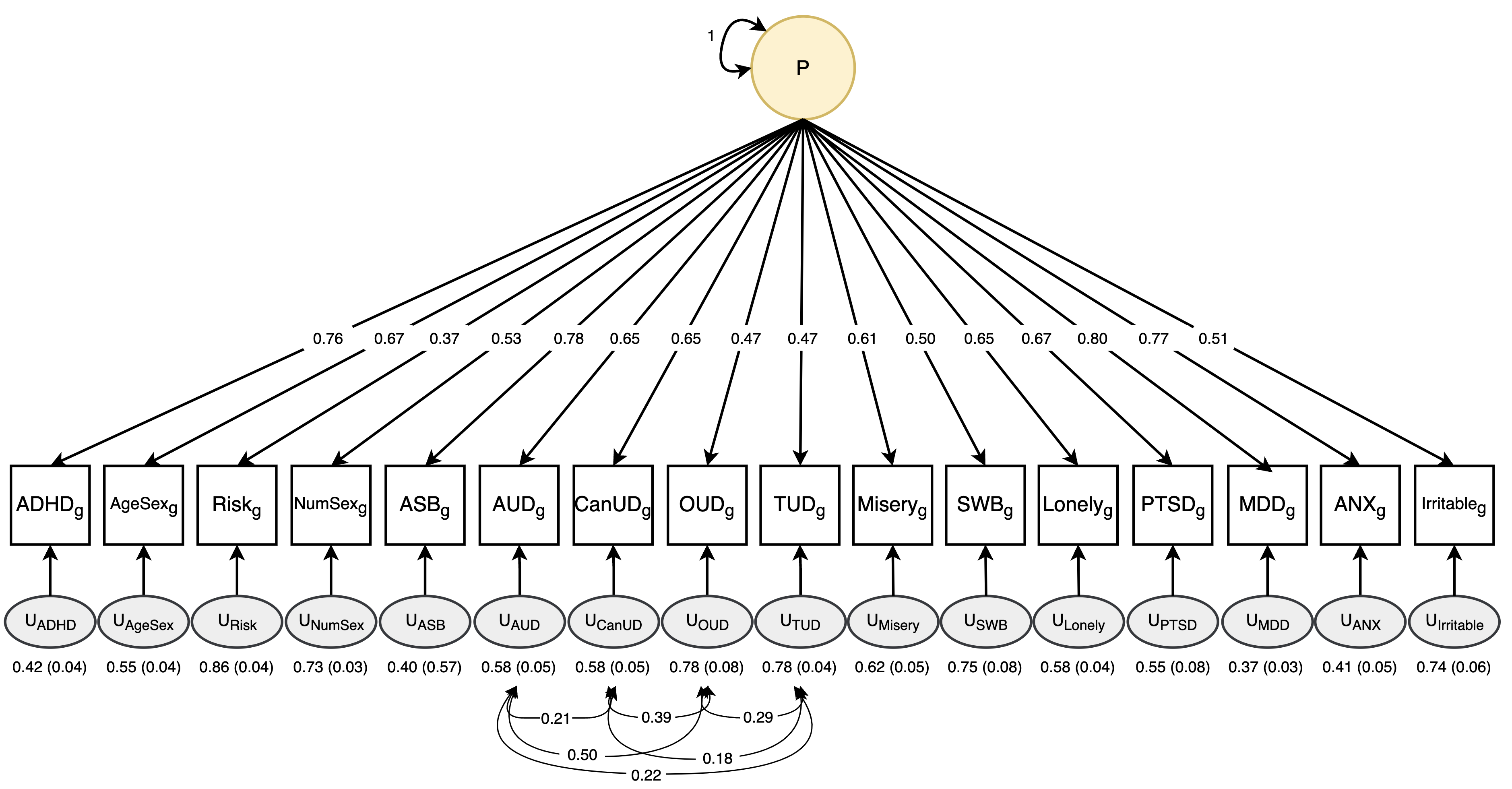

### **Supplementary Figure 2. General psychopathology confirmatory factor model.**

Model fit: 𝜒^2^(98) = 8965.28, p = 0, AIC = 9041.28, CFI = 0.79, SRMR = 0.15. P = general psychopathology, ADHD = attention deficit hyperactivity disorder, AgeSex = age at first sexual intercourse (reverse-coded), NumSex = number of sexual partners, ASB = antisocial behavior, AUD = alcohol use disorder, CanUD = cannabis use disorder, OUD = opioid use disorder, TUD = tobacco use disorder, SWB = subjective wellbeing (reverse-coded), PTSD = posttraumatic stress disorder, MDD = major depressive disorder, ANX = anxiety.

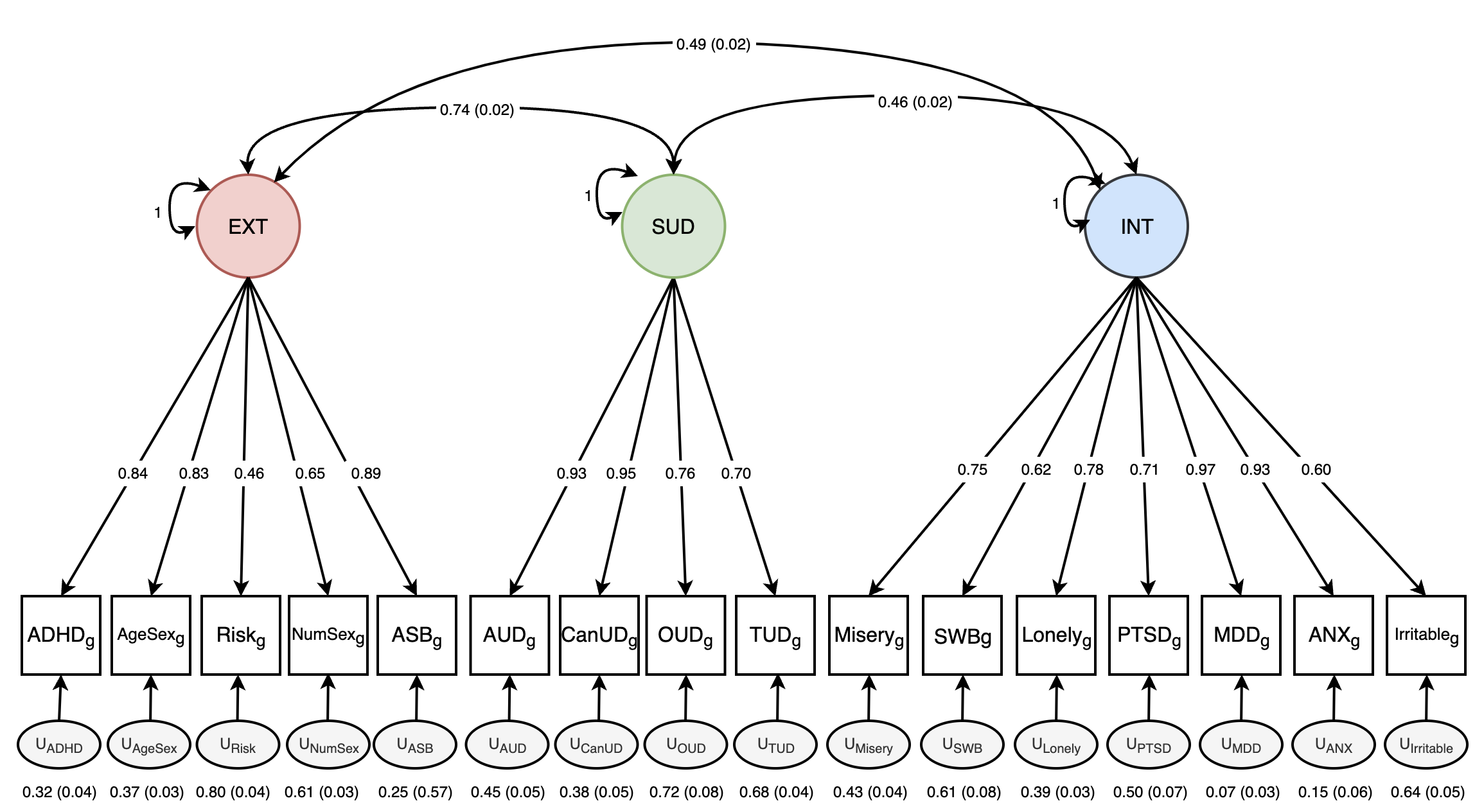

### **Supplementary Figure 3. Three correlated factors confirmatory factor model.**

Model fit: $\chi$^2^(101) = 4401.18, AIC = 4471.18, CFI = 0.90, and SRMR = 0.09. ADHD = attention deficit hyperactivity disorder, AgeSex = age at first sexual intercourse (reverse-coded), NumSex = number of sexual partners, ASB = antisocial behavior, AUD = alcohol use disorder, CanUD = cannabis use disorder, OUD = opioid use disorder, TUD = tobacco use disorder, SWB = subjective wellbeing (reverse-coded), PTSD = posttraumatic stress disorder, MDD = major depressive disorder, ANX = anxiety

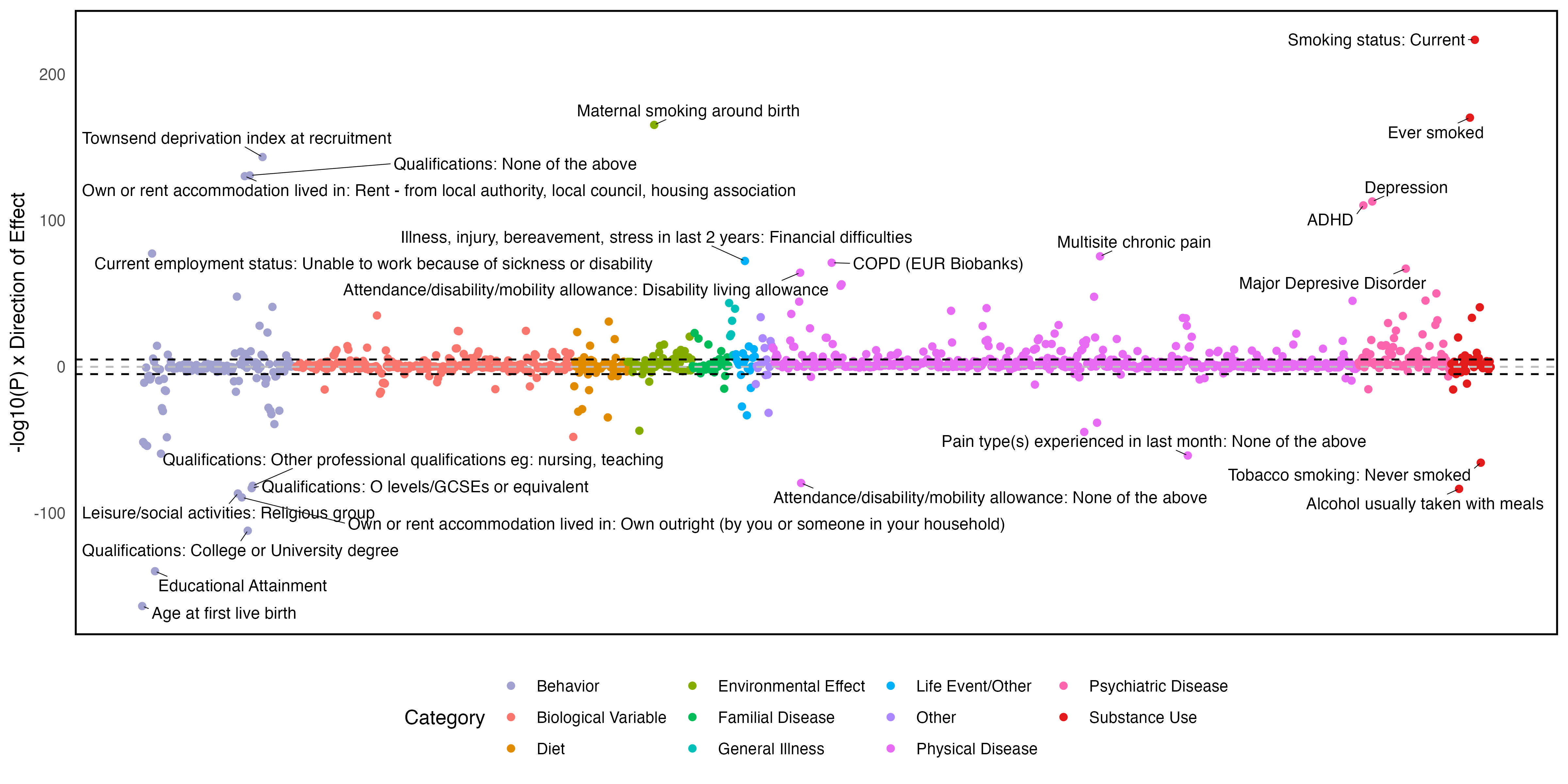

### **Supplementary Figure 4. Genetic correlations between the externalizing factor and publicly available phenotypes.**

The top 25 associations are annotated.

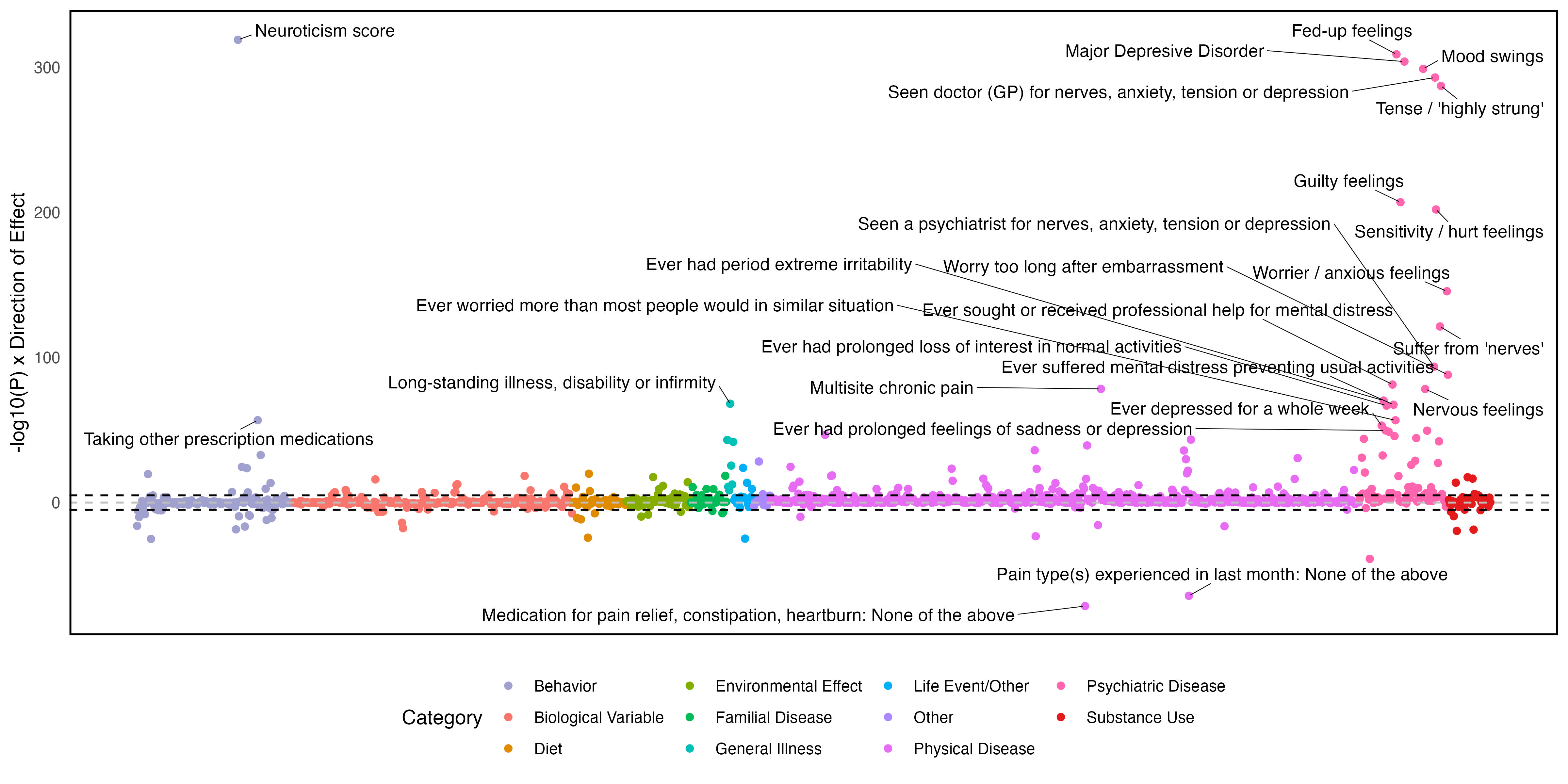

### **Supplementary Figure 5. Genetic correlations between the internalizing factor and publicly available phenotypes.**

The top 25 associations are annotated.

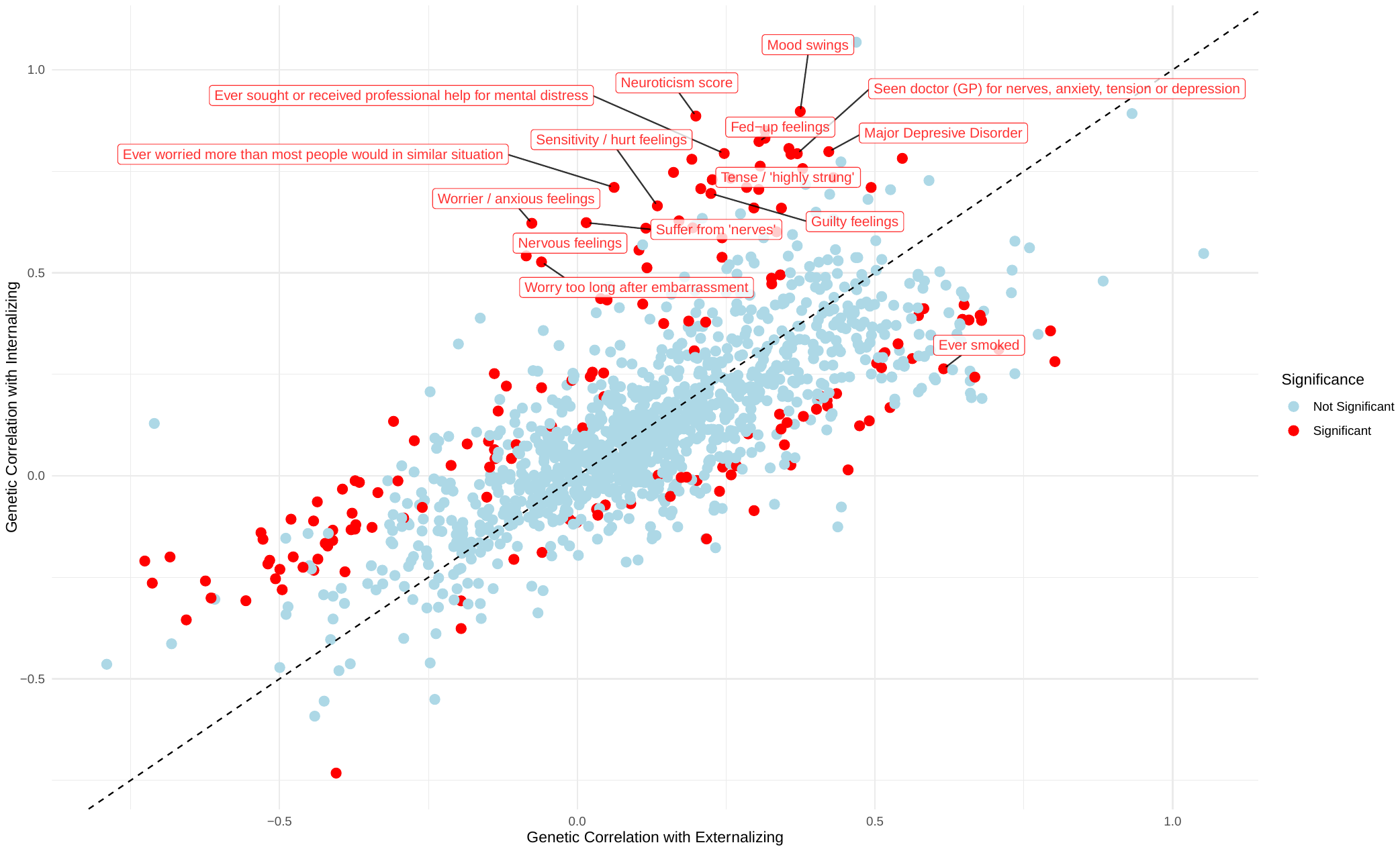

### **Supplementary Figure 6. Scatterplot of the genetic correlations for externalizing and internalizing.**

Red indicates significantly different genetic correlations after false discovery rate correction, and the genetic correlations with the greatest differences across spectra are annotated.
